# Post SARS-CoV-2 cell mediated Immune profiles; Case studies

**DOI:** 10.1101/2022.07.02.22276875

**Authors:** Rajveer Singh, Velayutham Ravichandiran, Dipanjan Ghosh, Ujjaini Ray, Sarbani Dasgupta, Soma Dutta, Aditi Saha, Debabrota Roy, Srinika Ghosh, A. Somasundaram, Pallab Dutta, N K Ganguly, Debatosh Datta

## Abstract

Cell-mediated immunity (CMI), which includes T-cells (both T helper and cytotoxic), is critical for effective antiviral defenses against coronavirus disease-2019 (COVID-19). To better understand the immunological characteristics of CD markers on T-cells in post-COVID-19 patients, we investigated the expression of differential CD markers in the patient groups in this study.

Flow cytometry was used to quantify total lymphocyte count and assess the levels of expression of CD markers in the samples. The percentage of Lymphocytes decreased significantly in the post-SARS-COV-2 patients in comparison to normal subjects, which is usually happening in any viral infection. In contrast to that, expression of CD8 was increased in the patient group having long SARS-COV-2 infection with comorbid complications with respect to the normal individuals and long SARS-COV-2 infection without comorbid complications. This data revealed that the cellular immunological responses corroborated with an earlier report of COVID-19 infection were mediated by CD8 upregulation and cytotoxic T lymphocyte hyperactivation.

## Introduction

Coronavirus Disease 2019 (COVID-19) is characterized by severe acute respiratory symptoms caused by a new beta coronavirus known as severe acute respiratory syndrome coronavirus-2 (SARS-CoV-2) and is the seventh coronavirus species to infect humans. SARS-CoV-2 is a member of the Nidovirales virus family, including the Coronaviridae [1]. Symptomatic transmission with symptoms ranging from severe form (shortness of breath, fever, and dry cough) to mild form (fever and dry cough). Blood oxygen saturation cut-off is 93 percent, and respiration frequency is greater than 30/min [2]. SARS-CoV-2 has been identified as an enveloped positive-sense single-stranded RNA virus (+ssRNA) with three and five caps. SARS-CoV-2 is distinguished from other RNA viruses by a particular furin location in the spike protein, as well as the mutation-prone and phylogenetically messy open reading frame1ab (Orf1ab) [3]. SARSCoV-2 is a round or elliptic, frequently pleomorphic virus with a diameter of 60–140 nm that is susceptible to UV radiation and heat [4, 5]. Spike (S), nucleocapsid (N), membrane (M), and envelope (E) are the major components of the entire structure (E), having a genome size of 30kb [6]. Angiotensin-converting enzyme 2 (ACE2), which is located on the surface of lung alveolar epithelial cells and small intestine enterocytes, has been proposed as the SARS-CoV-2 entrance site. Angiotensin II, a proinflammatory agent in the lungs, is broken down by ACE2. Inhibition of ACE2 could contribute to lung injury and the source of systemic inflammation with cytokine release, which can lead to ARDS and multiorgan failure. Some of the increased risks of unfavorable outcomes in people with COVID-19-related cardiovascular disease are likely due to disruption in immune system modulation, higher metabolic demand, and procoagulant activity (CVD). Myocardial damage, myocarditis, acute myocardial infarction, heart failure, dysrhythmias, and venous thromboembolic events are all cardiovascular system problems associated with SARS-COV-2 [7, 8]. The interconnections between the body’s innate and adaptive immune systems determine how the host reacts to viral infections. T lymphocytes, including CD4 T lymphocytes and CD8 T cells, play a crucial part in the immune system’s antiviral responses to viruses. Antiviral immune responses resulted in changes in the typical CD4:CD8 ratio in virally infected patients.

From previous studies, it has been known that, levels of intermediate CD16 LDNs (low density neutrophiles) are higher in severe patients than moderate patients, indicating that intermediate CD16 LDNs correlate with disease severity [9]. CD138 is another CD marker belonging to the syndecan family of type I transmembrane proteoglycans, which is made up of a core protein glycosylated with chondroitin and heparan sulfate (HS) moieties. CD138 has been linked to wound healing, cell adhesion, endocytosis, and the ability to translocate to the nucleus, among other things.[10]. Another CD marker, i.e., CD19, is a B cell marker; it’s been used to identify malignancies that originate from these cells, such as B cell lymphomas, acute lymphoblastic leukemia (ALL), and chronic lymphocytic leukemia (CLL) (CLL). CD19 is expressed at normal to high levels in the majority of B cell malignancies [11]. CD56 is the typical phenotypic marker of natural killer cells. However, it can also be seen in alpha/beta T cells, gamma/delta T cells, dendritic cells, and monocytes [12].

## Material and Methods

### Reagents and Material

CDs markers antibodies were purchased from BD Bioscience, India with following catalog numbers CD4 (561005), CD8 (561952), CD16 (560995), CD56 (340363), CD69(560968), CD138 (347193), CD45 (340953), and CD19 (340409).

### Clinical samples

Clinical samples were collected from Apollo multispeciality. Patients were grouped as-

1. Normal Subjects- (Figure S1).
2. Current SARS-COV-2 infection with Cardiac–Coronary presentation (Figure S3).
3. No SARS-COV-2 infection with Cardiac--Coronary presentation (Figure S4).
4. Recent SARS-COV-2 infection with Cardiac–Coronary presentation (Figure S5).
5. Long SARS-COV-2 infection with Cardiac—Coronary presentation (Figure S2).
6. Long SARS-COV-2 infection with Renal presentation (Figure S6).
7. Long SARS-COV-2 infection with No Clinical Presentation (Figure S7).

Usually, long covid cases are included in the group subjected to the time window of 3-4 months. However, this patient presented overt cardiac presentation after 22 months, without having any prior cardiac indication, even to a mild extent. Fresh whole blood of 5ml samples was collected from patients belonging to the above-described studied groups in EDTA anticoagulant tubes.

### Quantification of CD markers

100 μl of blood sample were added to the FACS tube. Further, 5 μl of each antibody per 100 μl of blood sample were added. Samples were incubated for 15min. Next,3ml of 1X lysis buffer was added following 10 min incubation. Samples were then centrifuged at 1500 rpm for 5min. Further, the supernatant was discarded, and the cell pellet was resuspended in 1X PBS. Samples were centrifuged again at 1500 rpm for 5min. The supernatant was discarded, and 1X FBS was added. CD markers were quantified by BD LSR II Fortessa Flow Cytometer on the sampling day and analyzed with Flowjo software.

## Results

### Clinical characteristics of study subjects

Essence of the study lies in determining the expression of CDs marker in post-COVID-19 coronary and renal presentation compared with normal subjects.

As anticipated, according to the covid-19 severity with the cardiac presentation, the clinical subsets were characterized by distinct changes in the expression of immune biomarkers and lymphocyte subsets.

### Immune biomarker expression

Total lymphocyte count was higher in the normal subjects/uncomplicated post-covid-19 patient compared to the patient having long/post covid-19 coronary presentation.

Patients were grouped as given in material and method. Patients having long SARS COV 2 infection with coronary presentation had significantly higher levels of CD8 as compared with others. Elevated CD4 levels in long SARS-COV-2 infection patients without complication compared to the normal subjects. Decrease in CD19 and an increase in CD8 in long SARS-COV-2 infection having renal complication was found compared to normal individuals and patients with SARS COV 2 infection without complications. One patient (TC), with a cardiac presentation, without clinically apparent SARS-COV-2 infection, showed a high CS8 population. This needs further intention in any subsequent prolonged study.

Active SARS COV 2 patient having cardiac presentation showed higher levels of CD19 compared to other groups. Long infection with SARS-COV-2 showed an increased level of CD4, and CD8. Active SARS-COV-2 infection increased level of CD19. Presence of comorbid complications showed variation in levels of immune biomarkers in patients, as shown in table 1.

**Table 1.**
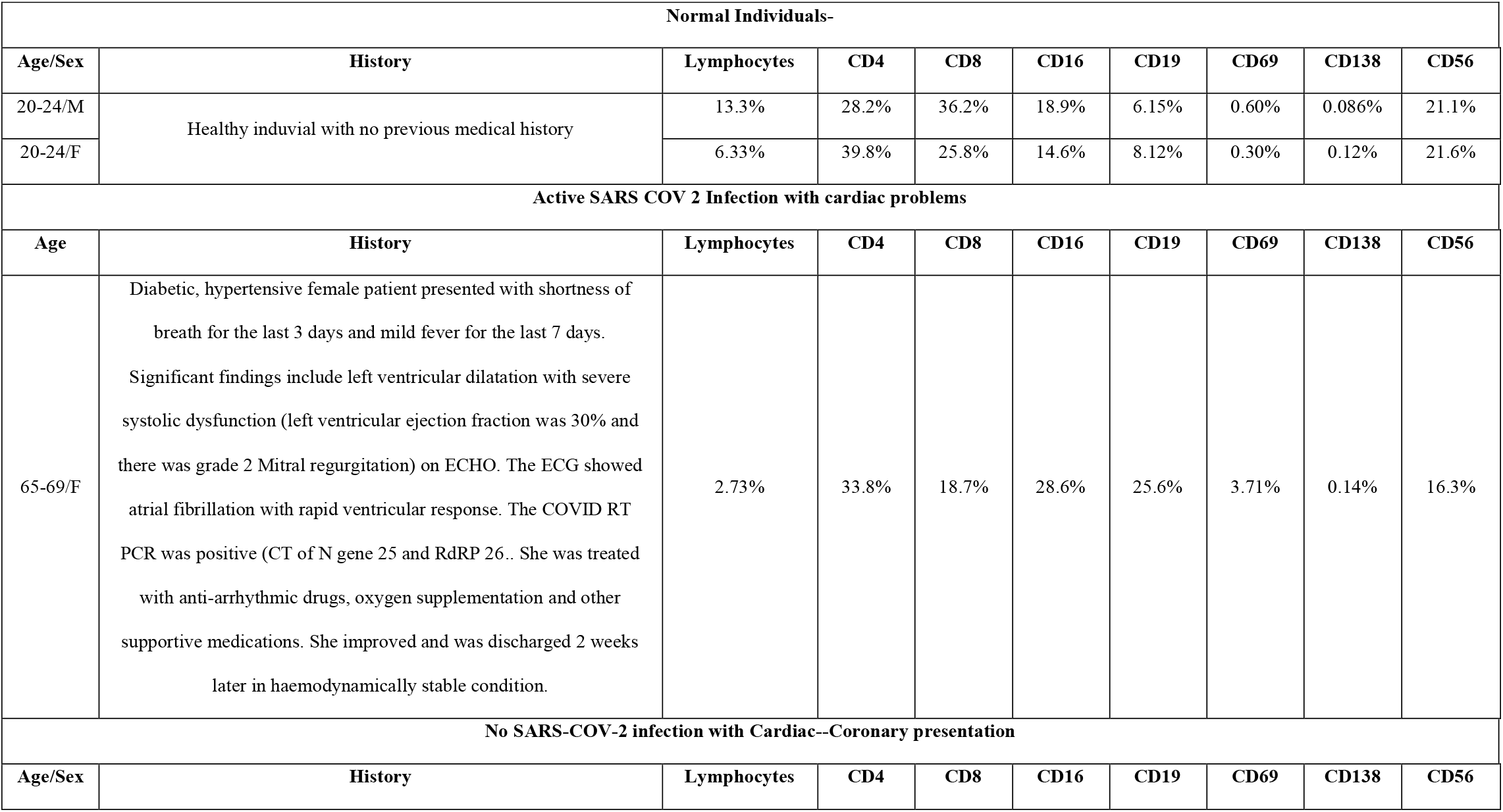
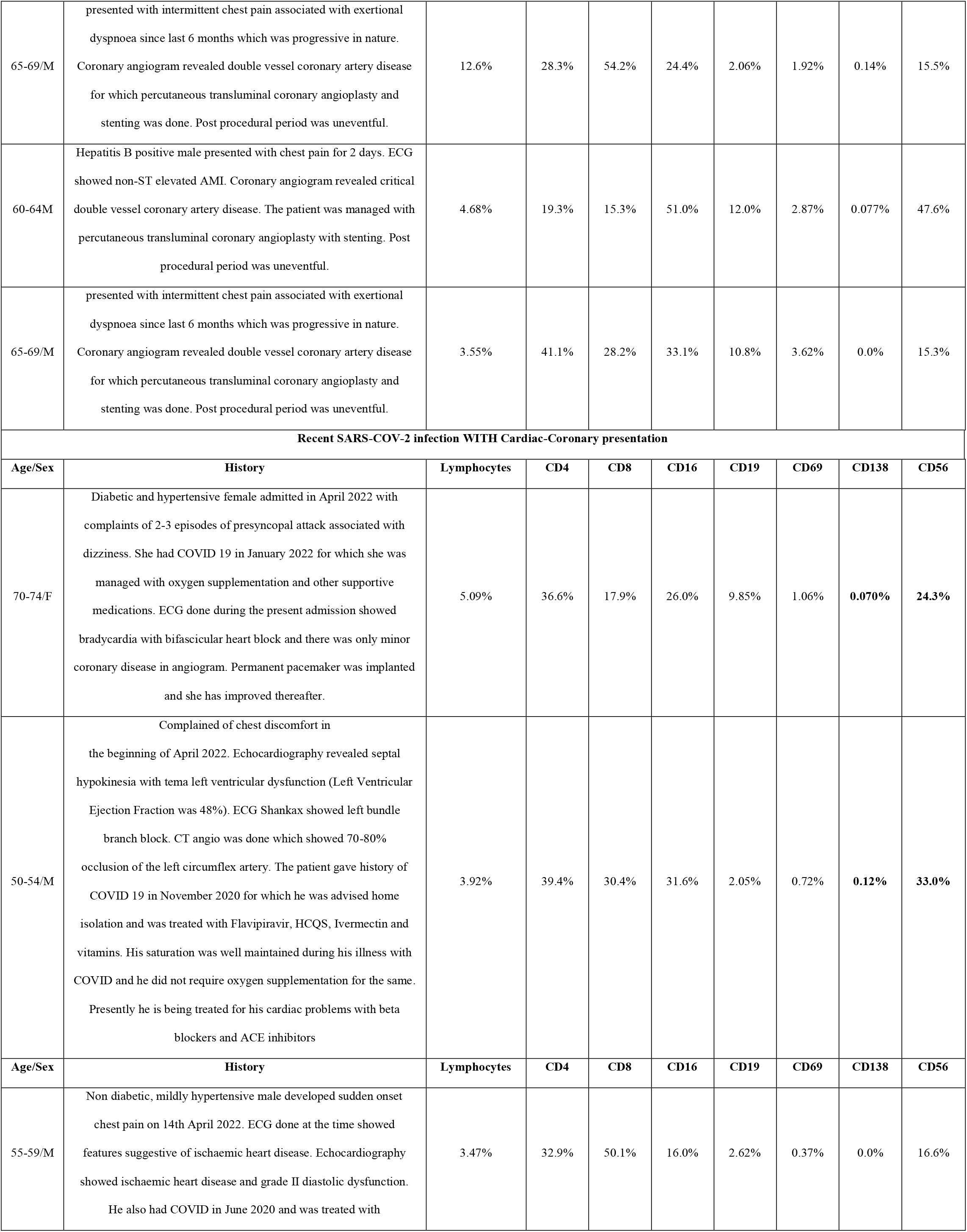
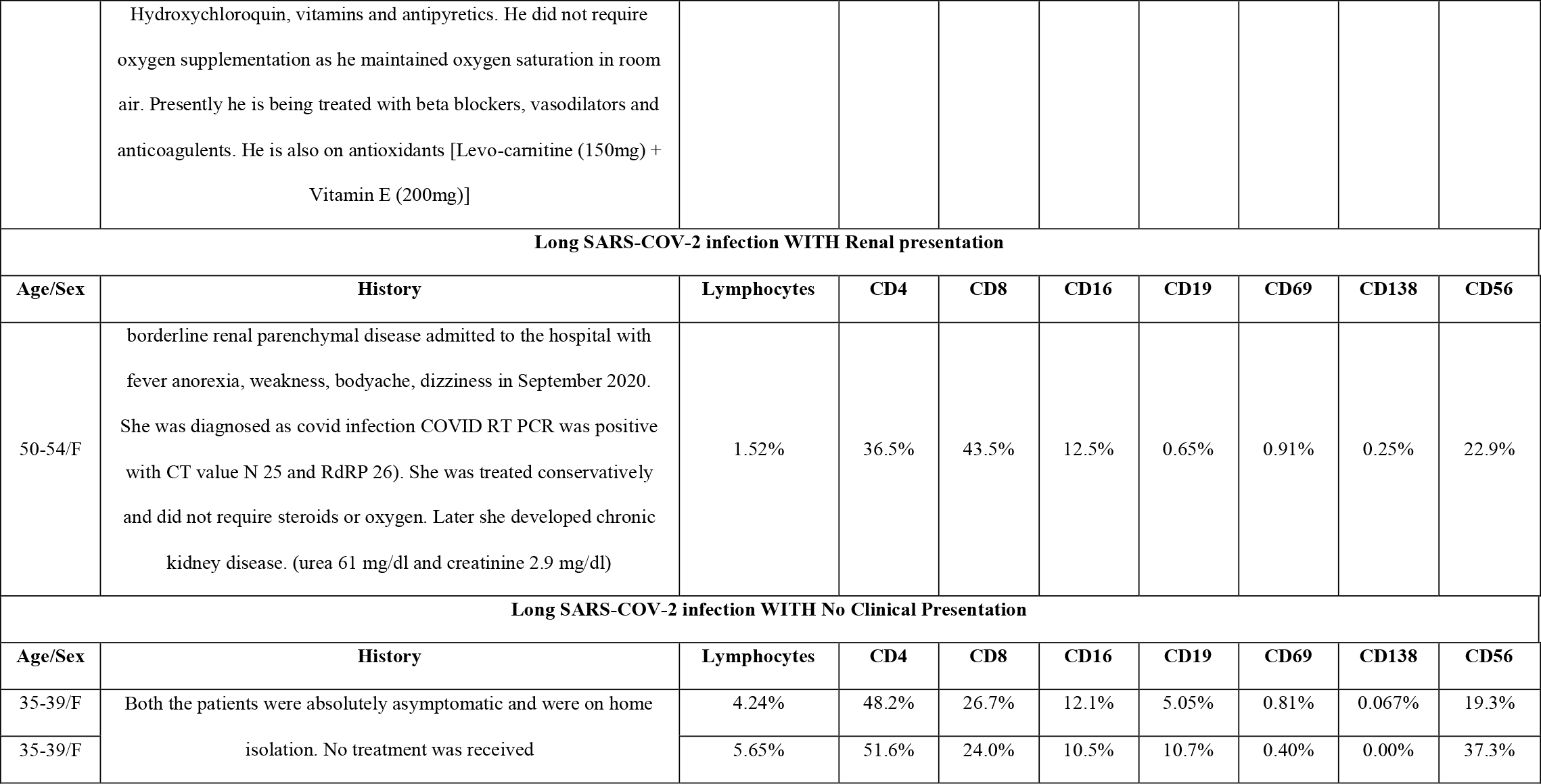
Represents the Patient’s immune biomarker expression.

| Normal Individuals- |  |  |  |  |  |  |  |  |  |
| --- | --- | --- | --- | --- | --- | --- | --- | --- | --- |
| Age/Sex | History | Lymphocytes | CD4 | CD8 | CD16 | CD19 | CD69 | CD138 | CD56 |
| 20-24/M | Healthy indivual with no previous medical history | 13.3% | 28.2% | 36.2% | 18.9% | 6.15% | 0.60% | 0.086% | 21.1% |
| 20-24/F |  | 6.33% | 39.8% | 25.8% | 14.6% | 8.12% | 0.30% | 0.12% | 21.6% |
| Active SARS COV 2 Infection with cardiac problems |  |  |  |  |  |  |  |  |  |
| Age | History | Lymphocytes | CD4 | CD8 | CD16 | CD19 | CD69 | CD138 | CD56 |
| 65-69/F | Diabetic, hypertensive female patient presented with shortness of breath for the last 3 days and mild fever for the last 7 days.<br>Significant findings include left ventricular dilatation with severe systolic dysfunction (left ventricular ejection fraction was 30% and there was grade 2 Mitral regurgitation) on ECHO. The ECG showed atrial fibrillation with rapid ventricular response. The COVID RT PCR was positive (CT of N gene 25 and RdRP 26.. She was treated with anti-arrhythmic drugs, oxygen supplementation and other supportive medications. She improved and was discharged 2 weeks later in haemodynamically stable condition. | 2.73% | 33.8% | 18.7% | 28.6% | 25.6% | 3.71% | 0.14% | 16.3% |
| No SARS-COV-2 infection with Cardiac--Coronary presentation |  |  |  |  |  |  |  |  |  |
| Age/Sex | History | Lymphocytes | CD4 | CD8 | CD16 | CD19 | CD69 | CD138 | CD56 |
| 65-69/M | presented with intermittent chest pain associated with exertional dyspnoea since last 6 months which was progressive in nature. Coronary angiogram revealed double vessel coronary artery disease for which percutaneous transluminal coronary angioplasty and stenting was done. Post procedural period was uneventful. | 12.6% | 28.3% | 54.2% | 24.4% | 2.06% | 1.92% | 0.14% | 15.5% |
| 60-64M | Hepatitis B positive male presented with chest pain for 2 days. ECG showed non-ST elevated AMI. Coronary angiogram revealed critical double vessel coronary artery disease. The patient was managed with percutaneous transluminal coronary angioplasty with stenting. Post procedural period was uneventful. | 4.68% | 19.3% | 15.3% | 51.0% | 12.0% | 2.87% | 0.077% | 47.6% |
| 65-69/M | presented with intermittent chest pain associated with exertional dyspnoea since last 6 months which was progressive in nature. Coronary angiogram revealed double vessel coronary artery disease for which percutaneous transluminal coronary angioplasty and stenting was done. Post procedural period was uneventful. | 3.55% | 41.1% | 28.2% | 33.1% | 10.8% | 3.62% | 0.0% | 15.3% |
| **Recent SARS-COV-2 infection WITH Cardiac-Coronary presentation** |  |  |  |  |  |  |  |  |  |
| **Age/Sex** | **History** | **Lymphocytes** | **CD4** | **CD8** | **CD16** | **CD19** | **CD69** | **CD138** | **CD56** |
| 70-74/F | Diabetic and hypertensive female admitted in April 2022 with complaints of 2-3 episodes of presyncopal attack associated with dizziness. She had COVID 19 in January 2022 for which she was managed with oxygen supplementation and other supportive medications. ECG done during the present admission showed bradycardia with bifascicular heart block and there was only minor coronary disease in angiogram. Permanent pacemaker was implanted and she has improved thereafter. | 5.09% | 36.6% | 17.9% | 26.0% | 9.85% | 1.06% | **0.070%** | **24.3%** |
| 50-54/M | Complained of chest discomfort in the beginning of April 2022. Echocardiography revealed septal hypokinesia with left ventricular dysfunction (Left Ventricular Ejection Fraction was 48%). ECG showed left bundle branch block. CT angio was done which showed 70-80% occlusion of the left circumflex artery. The patient gave history of COVID 19 in November 2020 for which he was advised home isolation and was treated with Flavipiravir, HCQS, Ivermectin and vitamins. His saturation was well maintained during his illness with COVID and he did not require oxygen supplementation for the same. Presently he is being treated for his cardiac problems with beta blockers and ACE inhibitors | 3.92% | 39.4% | 30.4% | 31.6% | 2.05% | 0.72% | **0.12%** | **33.0%** |
| **Age/Sex** | **History** | **Lymphocytes** | **CD4** | **CD8** | **CD16** | **CD19** | **CD69** | **CD138** | **CD56** |
| 55-59/M | Non diabetic, mildly hypertensive male developed sudden onset chest pain on 14th April 2022. ECG done at the time showed features suggestive of ischaemic heart disease. Echocardiography showed ischaemic heart disease and grade II diastolic dysfunction. He also had COVID in June 2020 and was treated with | 3.47% | 32.9% | 50.1% | 16.0% | 2.62% | 0.37% | 0.0% | 16.6% |
|  | Hydroxychloroquin, vitamins and antipyretics. He did not require oxygen supplementation as he maintained oxygen saturation in room air. Presently he is being treated with beta blockers, vasodilators and anticoagulents. He is also on antioxidants [Levo-carnitine (150mg) + Vitamin E (200mg)] |  |  |  |  |  |  |  |  |
| **Long SARS-COV-2 infection WITH Renal presentation** |  |  |  |  |  |  |  |  |  |
| **Age/Sex** | **History** | **Lymphocytes** | **CD4** | **CD8** | **CD16** | **CD19** | **CD69** | **CD138** | **CD56** |
| 50-54/F | borderline renal parenchymal disease admitted to the hospital with fever anorexia, weakness, bodyache, dizziness in September 2020. She was diagnosed as covid infection COVID RT PCR was positive with CT value N 25 and RdRP 26). She was treated conservatively and did not require steroids or oxygen. Later she developed chronic kidney disease. (urea 61 mg/dl and creatinine 2.9 mg/dl) | 1.52% | 36.5% | 43.5% | 12.5% | 0.65% | 0.91% | 0.25% | 22.9% |
| **Long SARS-COV-2 infection WITH No Clinical Presentation** |  |  |  |  |  |  |  |  |  |
| **Age/Sex** | **History** | **Lymphocytes** | **CD4** | **CD8** | **CD16** | **CD19** | **CD69** | **CD138** | **CD56** |
| 35-39/F | Both the patients were absolutely asymptomatic and were on home isolation. No treatment was received | 4.24% | 48.2% | 26.7% | 12.1% | 5.05% | 0.81% | 0.067% | 19.3% |
| 35-39/F |  | 5.65% | 51.6% | 24.0% | 10.5% | 10.7% | 0.40% | 0.00% | 37.3% |

## Discussion

Innate and acquired immune systems are both activated in response to viral infections. Activation of the CMI, particularly T cell activation, is the effective memory population against viral infection, including SARS-COV-2 [13]. Antigen presentation pathway mediated by the major histocompatibility complex (MHC) class I plays a critical role in host defense against intracellular infections and cancer. MHC class I aids antiviral immunity by allowing viral antigens to be presented to CD8 cytotoxic T cells more easily. As a result, virus-infected cells are preferentially eliminated by activated CD8 cytotoxic T cells [14].

CD8 + cytotoxic T lymphocytes (CTLs) removes viruses from host by secreting a variety of chemicals such as perforin, granzyme, and interferons (IFNs) [13]. CD4 helper T cells (Th) assist cytotoxic T cells and B cells in eradicating viral particles [15]. Observation showed a remarkable decrease in the number of lymphocytes in the peripheral blood of SARS-COV-2 infected patients compared to normal subjects.

Furthermore, a substantial rise in the percentage of CD8 marker was observed in the patients with long SARS-COV-2 infection, having cardiac and renal presentation compared with control. Active SARS-COV-2 infection significantly increased the CD19 levels concerning control subjects. Immunological response of infected patients with COVID-19 requires a normal CD4:CD8 ratio. An increase in CD8 levels, as observed, led to an altered CD4:CD8 ratio, which is a hallmark of viral infection. T lymphocytes augment cytotoxic activity by raising the CD8 population [16]. This being a few case study compilations, there is a need to observe the differential expression of a CDs population as different variations/sub-variations in species keep arising in the environments. Immunological stories become further complicated with vaccination, incomplete and without vaccination. Cell-mediated immune study in different perceptions of society is a continuous need. As SARS-COV-2 is unlikely to leave the global sin any time soon. This research offered information on variations in immunologic markers in COVID-19 patients. The use of large populations and additional immunologic markers in such studies could pave the way for detecting and treating this condition. Apparently, post covid syndrome is restricted by a time window of 3-4 months. Given a patient profile and clinical presentation, it looks like there is a need to extend the time window to a very reasonable extent. The unpredictability of onset and immunological profile looks remarkable in a small number of patients presented here. There must be a common predictability/damaging marker in post-covid multiorgan infections. As per CMI concern, the CD4 population shows nearly no deviation from normal subjects. Whereas the CD8 population in long cardiac and renal cases significantly increases. There should be a standing question about affecting the memory population generated in the longer term. Post-vaccination IgG build-up goes significantly in a short period, although with a variation. An observed question should be (1) Covid vaccination, or active infection is longer memory development and a resident end point?. (2) Is the immunoglobulin decay period the total actual period that one remains covered through the humoral immune arm?. (3) CD8 is a cytotoxic cell population that probably will not give longer protection, and the scenario is further compounded by the rapid development of variation in viral species.

In conclusion, a study suggested a significant increase in the expression of CD8 in infection groups, inferring that the immune response to COVID-19 infection is mediated by CD8 upregulation and hyperactivation of Cytotoxic T-cells antiviral responses rather than by alterations in the CD4:CD8 ratio and CD4 percentage. In the respective type of vaccine, its boosting, over and above having an active infection is an in-effective inducer of memory/ short in memory.

## Data Availability

All data produced in the present work are contained in the manuscript

## Conflict of Interest

The author reports no conflict of interest

## Ethical statement

Ethical clearance was done, by ethical committee at Apollo multi-speciality hospital, Kolkata, following the ICMR guidelines.

---

**Figure S1.**
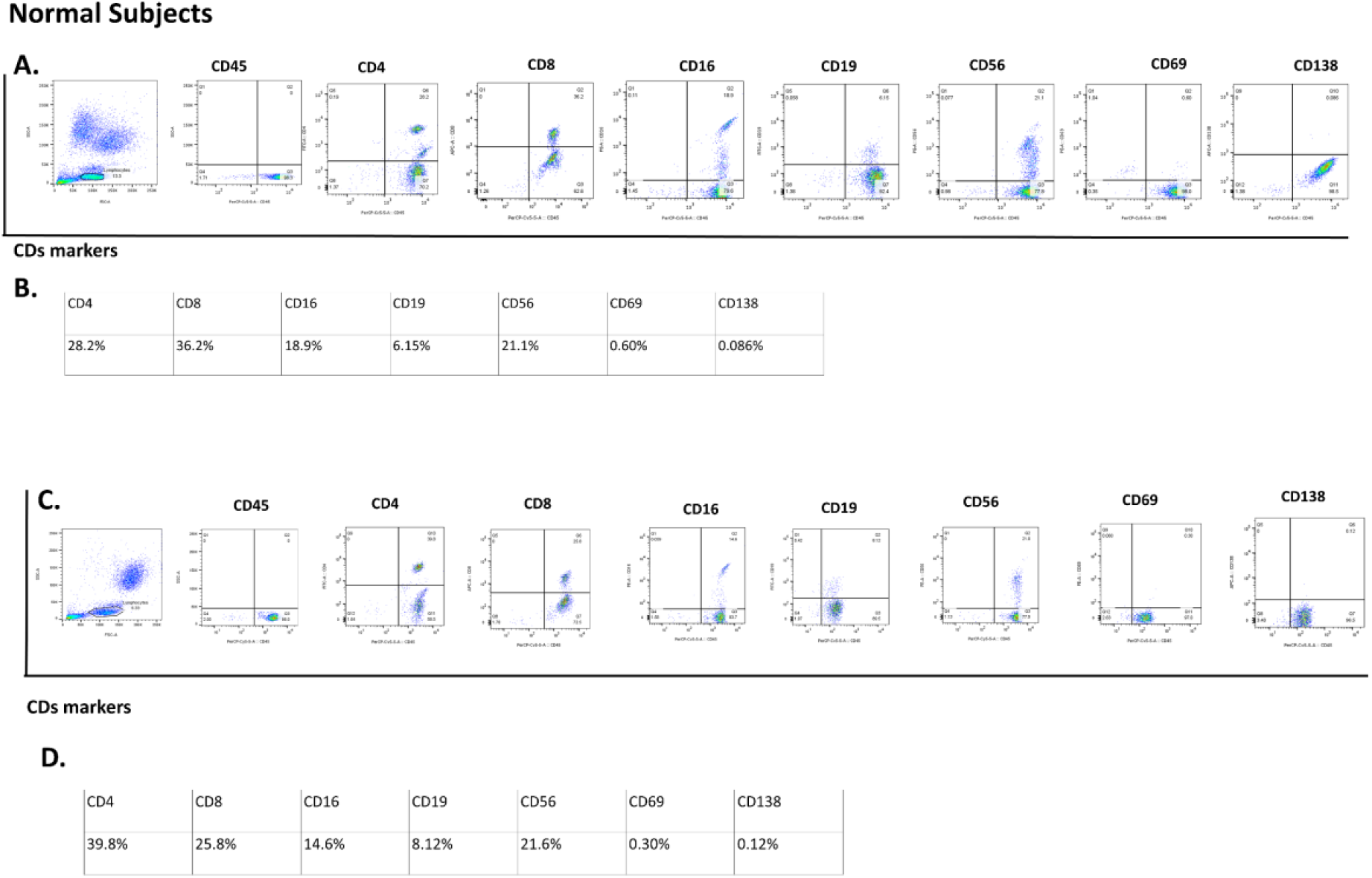
Represents the CDs marker screening of normal subjects. (A.), (C.) Quantification of CDs markers by flow cytometry of subject and (B), (D) represents the percentage of change in CDs expression.

**Figure S2.**
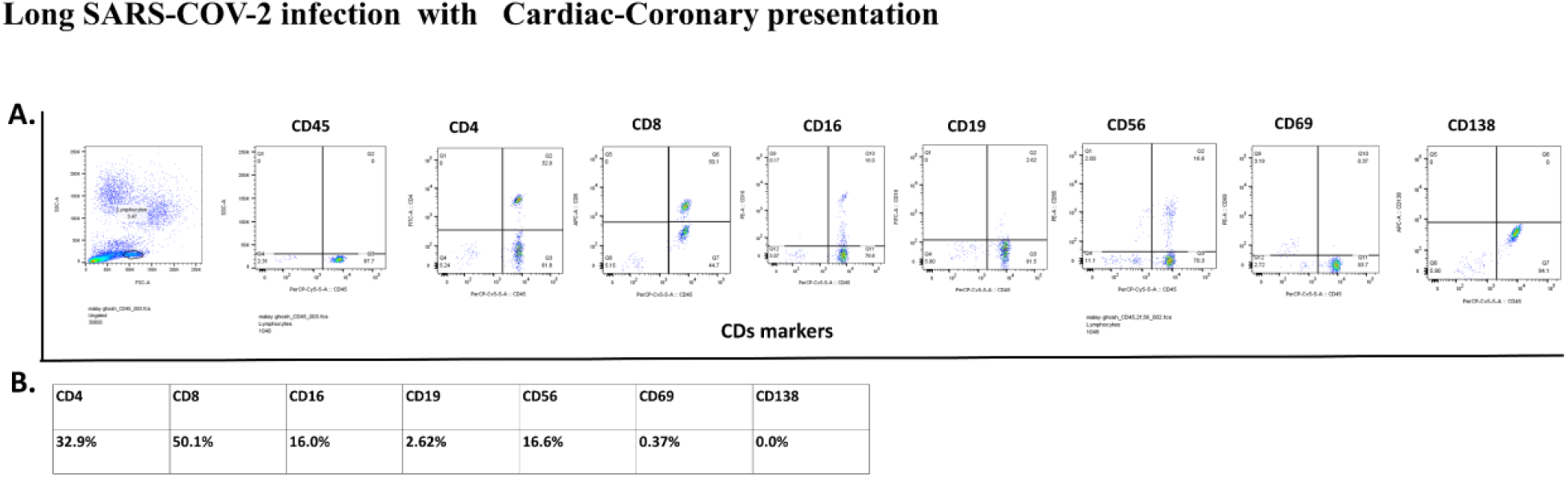
Represents the CDs marker screening of long SARS-COV-2 infection with cardiac-coronary presentation. (A) Quantification of CDs markers by flow cytometry, (B) Represents the percentage of change in CDs expression.

**Figure S3.**
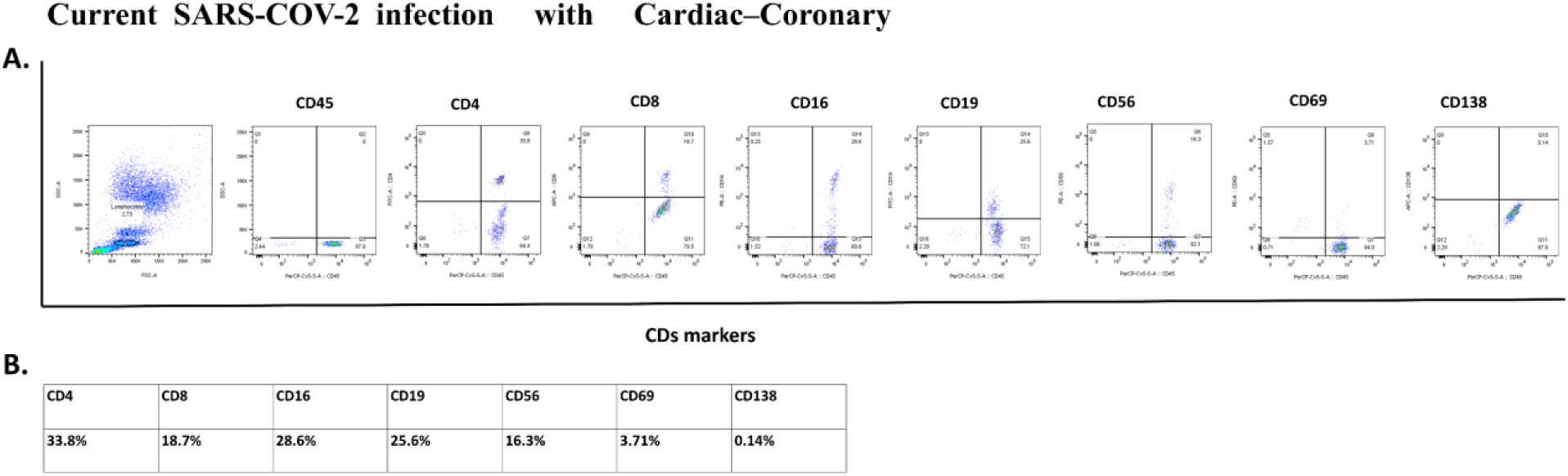
Represents the CDs marker screening current SARS-COV-2 infection with cardiac–coronary presentation. (A) Quantification of CDs markers by flow cytometry, and (B) Represents the percentage of change in CDs expression.

**Figure S4.**
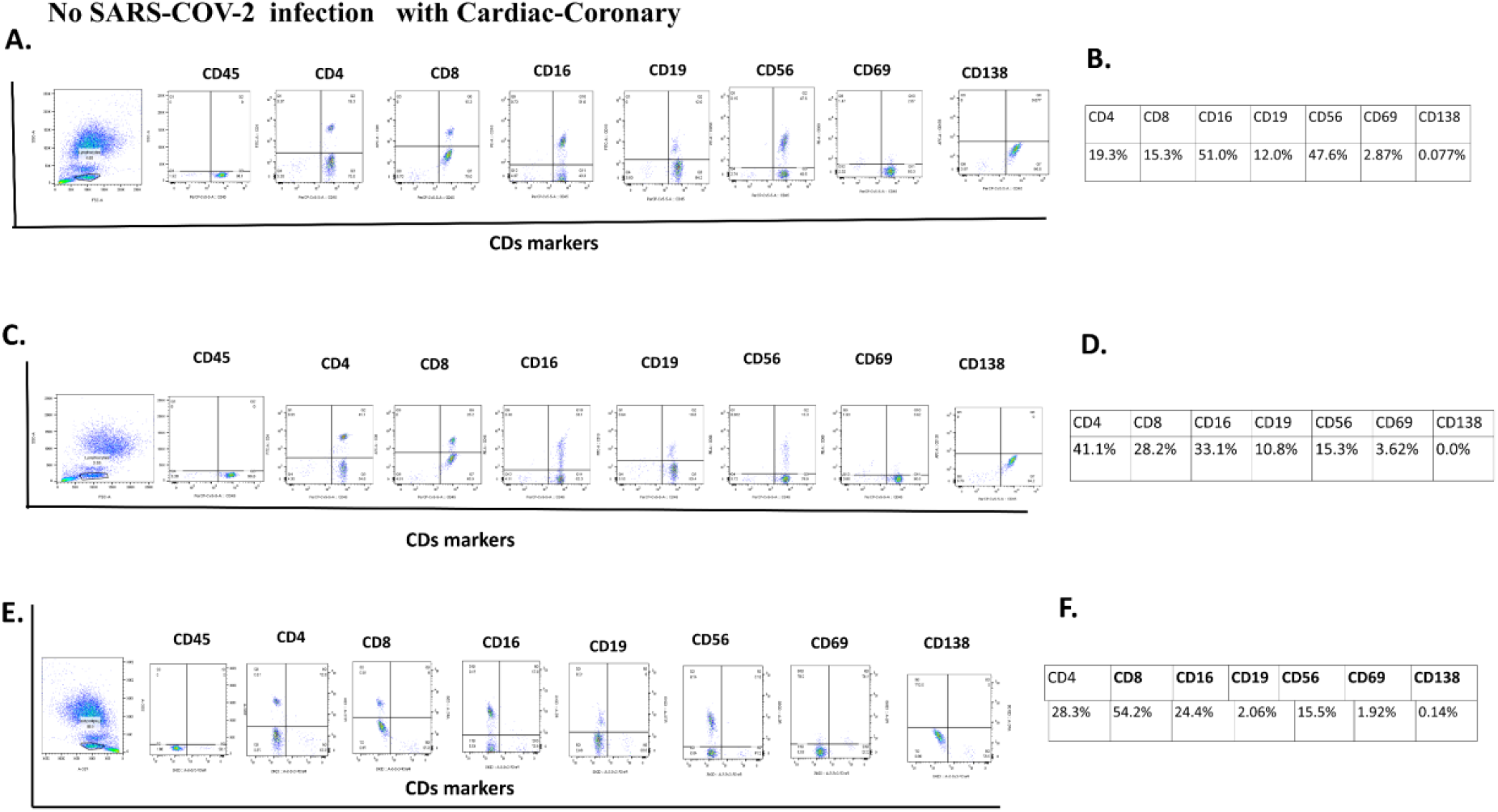
Represents the CDs marker screening of no SARS-COV-2 infection with Cardiac-Coronary presentation by flow cytometry (A), (C) and (E). And Represents the percentage of change in CDs expression (B), (D) and (E).

**Figure S5.**
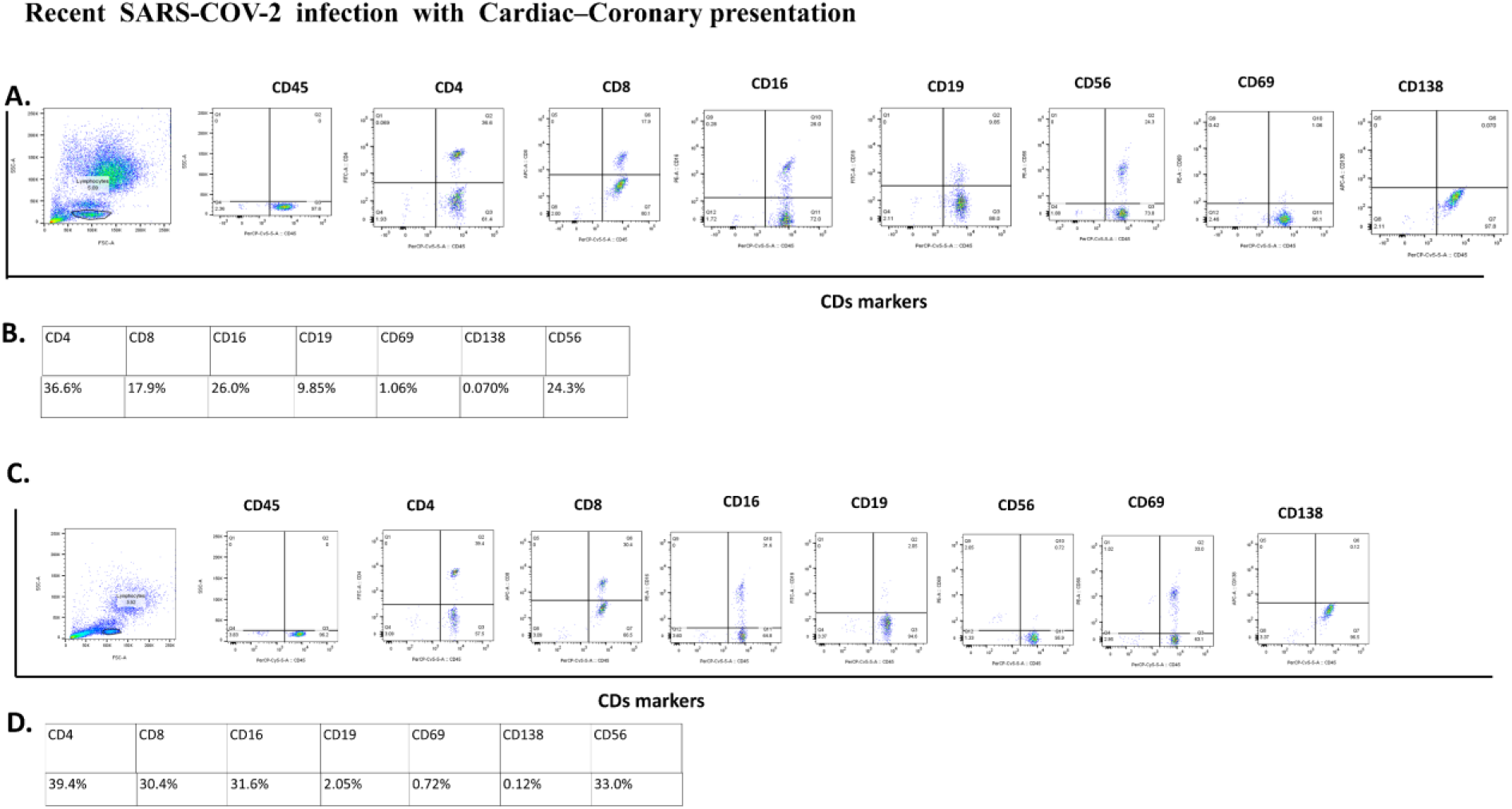
Represents the CDs marker screening of recent SARS-COV-2 infection with Cardiac–Coronary presentation. Quantification of CDs markers by flow cytometry (A)(D). Represents the percentage of change in CDs expression (B) and (D).

**Figure S6.**
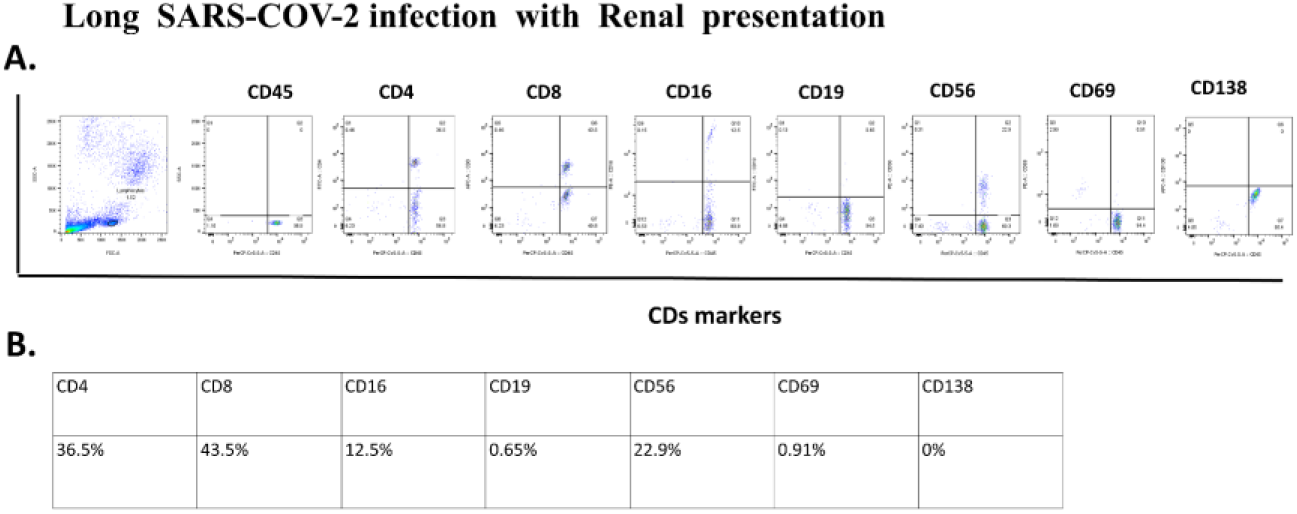
Represents the CDs marker screening of Long SARS-COV-2 infection with Renal presentation, (A) Quantification of CDs markers by flow cytometry, and (B) Represents the percentage of change in CDs expression.

**Figure S7.**
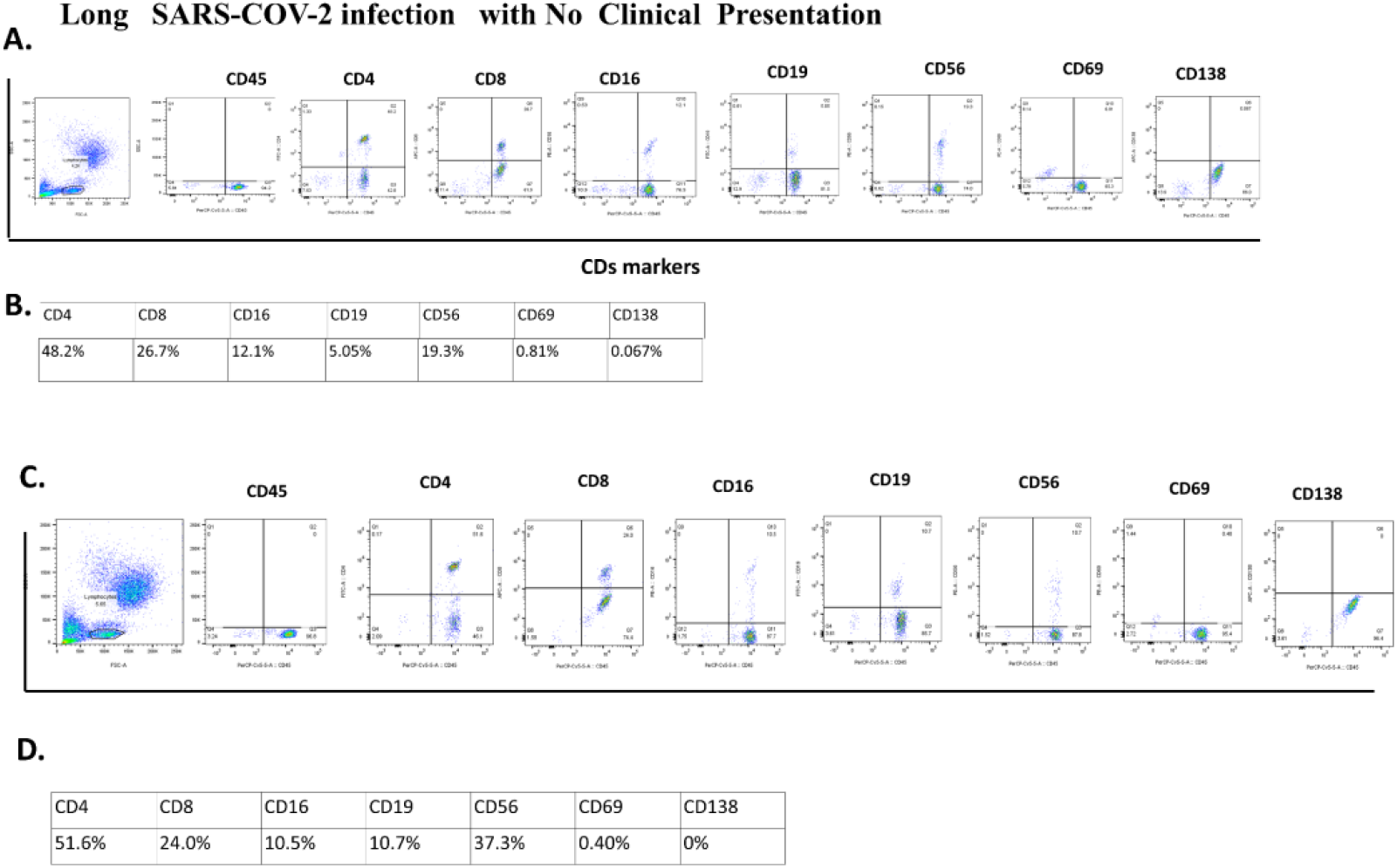
Represents the CDs marker screening of Long SARS-COV-2 infection with no clinical presentation Quantification of CDs markers by flow cytometry (A) (C). Represents the percentage of change in CDs expression (B) and (D).

## Notes

### Competing Interest Statement

The authors have declared no competing interest.

### Funding Statement

This study was funded by NIPER, Kolkata.

### Author Declarations

Ethical clearance was done, by ethical committee at Apollo multispeciality hospital, Kolkata, following the ICMR guidelines.

